# A Deep Learning Vision-Language Model for Diagnosing Pediatric Dental Diseases

**DOI:** 10.1101/2025.05.21.25328098

**Authors:** Tuan D. Pham

**Affiliations:** Barts and The London School of Medicine and Dentistry, Queen Mary University of London, Turner Street, London E1 2AD, United Kingdom

**Keywords:** Dental diseases, children, panoramic radiographs, deep learning, natural language processing, fuzzy recurrence plots, gray-level co-occurrence matrix

## Abstract

This study proposes a deep learning vision-language model for the automated diagnosis of pediatric dental diseases, with a focus on differentiating between caries and periapical infections. The model integrates visual features extracted from panoramic radiographs using methods of non-linear dynamics and textural encoding with textual descriptions generated by a large language model. These multimodal features are concatenated and used to train a 1D-CNN classifier. Experimental results demonstrate that the proposed model outperforms conventional convolutional neural networks and standalone language-based approaches, achieving high accuracy (90%), sensitivity (92%), precision (92%), and an AUC of 0.96. This work highlights the value of combining structured visual and textual representations in improving diagnostic accuracy and interpretability in dental radiology. The approach offers a promising direction for the development of context-aware, AI-assisted diagnostic tools in pediatric dental care.

## 1 Introduction

Pediatric dental diseases represent one of the most prevalent chronic conditions affecting children globally [1, 2, 3]. A systematic analysis estimated that oral conditions affected more than three and a half billion people worldwide in 2010, with untreated caries in primary teeth ranking among the ten most common conditions across all ages [4]. Early childhood caries alone can affect up to 70% of preschool-aged children in some regions [5], and up to 85% for disadvantaged groups [6], leading to pain, infection, and impaired nutrition and growth if left untreated. Beyond caries, developmental anomalies such as malocclusions [3], enamel anomalies [7], and periodontal conditions [8] further contribute to the disease burden in pediatric populations [9, 10, 11, 12].

Detecting these conditions at an early stage poses unique challenges. Children’s continuously changing dental anatomy and varying stages of tooth eruption can obscure radiographic signs of disease [13, 14]. Moreover, pediatric patients may have difficulty remaining still for high-quality radiographs, resulting in motion artifacts that complicate interpretation. Subtle lesions or early enamel demineralization often present with low contrast and can be easily overlooked, especially without standardized imaging protocols and in the absence of overt clinical symptoms [15].

Dental panoramic radiography is a cornerstone of pediatric dental assessment, offering a broad overview of both erupted and developing teeth, jawbone morphology, and surrounding structures in a single image [16, 17]. Clinicians typically interpret these images manually, combining visual inspection with clinical judgment to identify carious lesions, root abnormalities, and growth disturbances [18, 19]. While expert review remains the gold standard, it is inherently time-consuming and subject to variability.

Inter-observer variability in radiographic interpretation is well documented: one study reported significant discrepancies in the detection of proximal caries among examiners, with Cohen’s Kappa values ranging from fair to moderate agreement [20]. Such variability can lead to inconsistent diagnoses and treatment plans, undermining care quality. In addition, manual review of large imaging datasets imposes a substantial workload on dental practitioners, potentially delaying diagnosis and intervention. There is thus a clear need for automated, reproducible tools to support clinicians in the early detection and standardized classification of pediatric dental diseases.

Over the past decade, convolutional neural networks (CNNs), developed in artificial intelligence (AI), have become foundational to automated medical image analysis, achieving human-level performance in diverse tasks such as lesion detection, segmentation, and classification. In radiology, CNN-based systems have demonstrated high sensitivity and specificity for identifying pathologies on chest radiographs and CT scans, often rivaling expert radiologists [21]. Dermatology has similarly benefited: CNNs trained on clinical images accurately classify melanoma versus benign nevi, facilitating early skin cancer detection [22]. Recent dentistry research has increasingly leveraged AI tools to support various aspects of pediatric dental care, including dental development [23], pedodontics [24], caries detection [25], root fracture identification [26], and the localization of anatomical landmarks in intraoral and panoramic radiographs of children [27]. Additional studies have explored AI applications more broadly in pediatric dentistry [28, 29, 30].

More recently, transformer architectures, which were originally developed for natural language processing, have been adapted for vision tasks. Vision transformers (ViTs) segment images into patches and apply self-attention mechanisms, enabling the capture of long-range dependencies without convolutional inductive biases. ViTs have matched or surpassed CNN performance on large-scale benchmarks, and in medical imaging, they have shown promise for tasks requiring context-aware feature extraction, such as tumor segmentation in magnetic resonance imaging [31] and bone age assessment from hand radiographs [32]. Hybrid CNN–transformer models further combine localized feature learning with global context, improving robustness in settings with limited annotated data.

Vision-language models (VLMs) represent a growing frontier in medical AI, jointly learning visual and textual modalities to enhance classification, interpretability, and multimodal understanding. Pretrained large-scale VLMs such as CLIP leverage paired image–caption datasets to align visual embeddings with semantic text representations, yielding generalizable features applicable across downstream tasks [33]. In medicine, VLMs facilitate automatic radiology report generation: models trained on chest X-ray images and corresponding narrative reports can produce coherent, clinically relevant captions that assist radiologists in reporting workflows [34, 35, 36].

Image captioning and cross-modal retrieval further illustrate VLM utility. Captioning systems generate structured descriptions of visible findings, offering explainable insights into model decision-making [37, 38]. Cross-modal retrieval enables clinicians to query large image databases using text prompts, retrieving similar cases to inform diagnosis and treatment planning [39, 40]. By integrating visual cues with natural language, VLMs not only improve classification accuracy but also provide transparent, human-interpretable outputs that bridge the gap between algorithmic predictions and clinical reasoning.

Despite the rapid adoption of deep learning in dental imaging, the vast majority of studies have focused solely on image-based CNNs. Recent reviews highlight that CNNs dominate applications such as caries detection, anatomical landmark identification, and automated cephalometric analysis, achieving high accuracy but relying exclusively on pixel-based features [41, 42]. In contrast, VLMs, which have shown considerable promise in other medical domains, remain almost entirely unexplored in dentistry.

Even within the broader field of dental AI, there is a conspicuous lack of multimodal approaches that integrate both visual and textual information. This gap is particularly pronounced in pediatric dentistry, where anatomical variability and the evolving nature of dentition pose additional challenges. To date, no published studies have leveraged VLM architectures to jointly learn from panoramic radiographs and corresponding text descriptions or diagnostic labels in children, leaving a critical opportunity unaddressed.

Combining image data with textual interpretations can offer several advantages that can directly address the challenges of pediatric dental diagnosis. Multimodal models can learn complementary representations: visual encoders capture spatial and morphological features, while language encoders contextualize these features in clinically meaningful terms. This integration enhances robustness against artefacts and anatomical variability by grounding image-based predictions in semantic descriptions. Furthermore, the natural-language outputs of VLMs can be expected to improve explainability, enabling clinicians to verify that diagnostic cues align with their clinical reasoning. Such transparency is essential in pediatric care, where treatment decisions must carefully balance intervention benefits against developmental considerations. By addressing this gap, VLMs have the potential to elevate pediatric dental diagnostics beyond the capabilities of image-only CNNs, delivering standardized, interpretable, and resilient decision support tailored to pediatric patients.

The primary aim of this study is to develop and evaluate a VLM specifically tailored for the deep learning–based classification of pediatric dental diseases. Leveraging panoramic radiographs as the visual modality and clinically informed text descriptions as the linguistic modality, this approach seeks to harness the complementary strengths of both image and language representations. By integrating these modalities, the model is designed to not only assign accurate disease labels, but also to generate brief, human-interpretable summaries of the key radiographic findings that underlie each classification decision. The main contributions are outlined as follows.

### Hybrid Multimodal Architecture

A hybrid framework is introduced to combines a convolutional encoder with a transformer-based language decoder. This design enables joint learning from image features and text embeddings, allowing the model to align radiographic patterns with semantic descriptions.

### Clinically Interpretable Text Generation

The model produces expert-style text describing observed abnormalities, enhancing transparency and facilitating clinician review of automated predictions.

### Comparative Analysis with Baselines

To quantify the benefits of multimodal learning, a comparison against two baselines is performed: an image-only CNN model and a text-only transformer model to confirm if the vision-language approach can outperforms both baselines in classification accuracy.

The remainder of this paper is organized as follows. Section 2 describes the image and text datasets. Section 3 presents the AI models used for disease classification. Results are reported in Section 4. Section 5 discusses the implications of findings for pediatric dental practice and outlines directions for future research. Finally, Section 6 provides the concluding remarks of the study.

## 2 Data

### 2.1 Panoramic radiographs

A dental panoramic radiograph, also referred to as an orthopantomogram (OPG), is a two-dimensional dental imaging technique that captures a comprehensive view of the entire mouth in a single image [43]. This includes all the teeth in both the upper and lower jaws, as well as surrounding anatomical structures such as the alveolar bone, maxillary sinuses, temporo-mandibular joints, and portions of the nasal cavity [44]. By providing a broad overview of the oral and maxillofacial region, panoramic radiographs are instrumental in the diagnosis of a wide range of conditions, including impacted teeth, jaw fractures, cysts, tumors, periodontal disease, and temporomandibular joint disorders. They also play a useful role in treatment planning for orthodontic procedures, dental implants, extractions, and other surgical interventions.

The *Children’s Dental Panoramic Radiographs Dataset for Caries Segmentation and Dental Disease Detection* [45] is a publicly available dataset designed to support the development of deep learning models for pediatric dental diagnostics. It consists of panoramic radiographs collected from 106 pediatric patients aged between 2 and 13 years at Hangzhou Xiasha District Dental Hospital. Each image is accompanied by expert-generated annotations of tooth structures and dental diseases, created using EISeg for segmentation and LabelMe for disease labeling. Seven licensed dentists were involved in the annotation process to ensure clinical accuracy and consistency.

The dataset is structured into two main components: the segmentation dataset, which includes the original radiographs and corresponding binary masks outlining tooth structures; and the detection dataset, which contains the original images along with JSON files that specify disease annotations such as caries, pulpitis, and periodontitis. The release includes 100 core annotated pediatric images. The full dataset is available via *figshare* [46].

To enable comparison with a previous study [47], only the caries and periapical infection classes were selected, as they offer a balanced sample size suitable for machine learning classification. The dataset includes 29 images for each disease category, making a total of 58 panoramic radiographs. Figure 1 shows examples of pediatric dental panoramic radiographs of carries and periapical infections, which are considered in this study.

**Figure 1:**
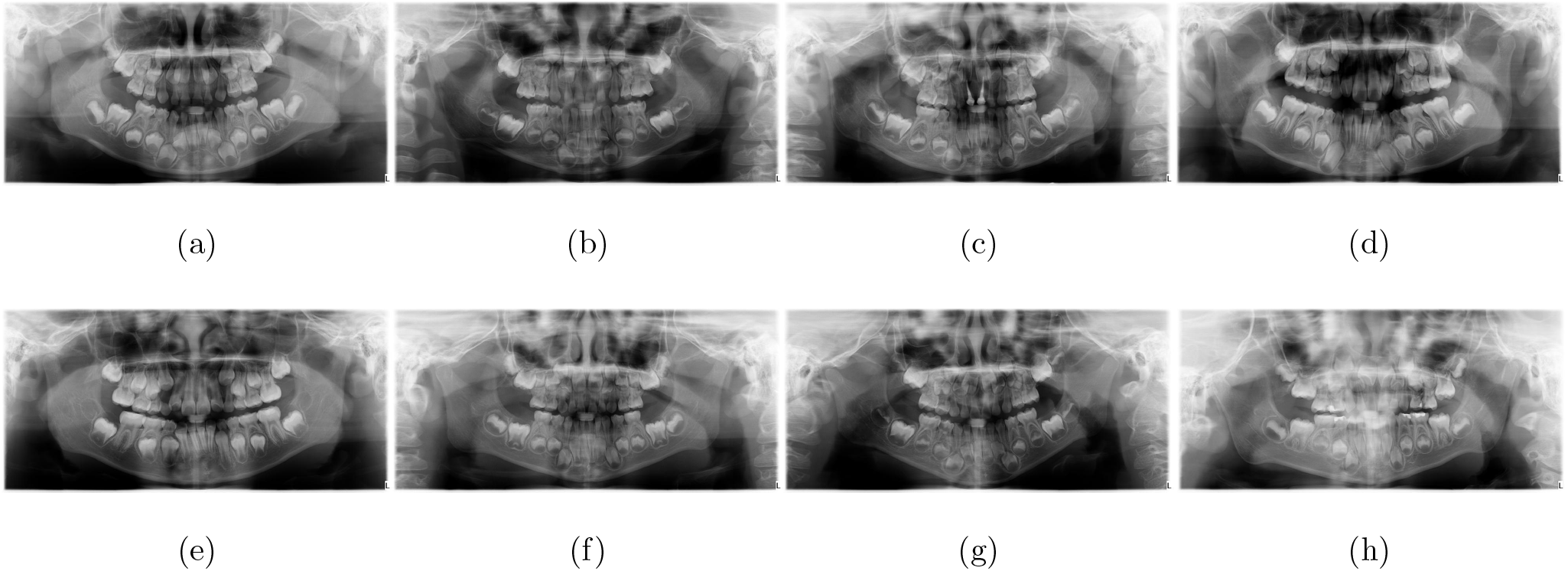
Pediatric dental panoramic radiograph of caries (a)-(d), and periapical infections (e)-(h).

### 2.2 Narratives of panoramic radiographs

The original panoramic radiographs were significantly downscaled, with their resolution reduced by a factor of 25 [47]. This resolution reduction was implemented primarily to enhance computational efficiency. High-resolution panoramic radiographs require considerable processing power, which can slow down image analysis and increase the time needed for processing large datasets. By reducing the resolution, the images could be analyzed more quickly while still retaining the essential features relevant for disease classification. Additionally, lower-resolution images require less storage space, further optimizing the performance of the analysis system.

Following this reduction, the images were processed using ChatGPT, powered by OpenAI’s advanced GPT-4 architecture [48]. ChatGPT was employed to generate concise, informative descriptions of potential pediatric dental diseases visible within the low-resolution images. Each description was carefully crafted to highlight key features indicative of specific diseases in approximately 12 words. This approach enabled a more efficient and streamlined classification process, focusing on the identification of relevant dental abnormalities or conditions.

ChatGPT has been reported as a useful AI tool in dentistry [49, 50, 51]. In the context of pediatric dental disease classification, its use for generating text descriptions offers several key benefits. First, the AI’s natural language processing capabilities allowed for the generation of precise, contextually relevant descriptions, improving the efficiency and accuracy of image interpretation. ChatGPT’s ability to generate consistent, unbiased descriptions also minimized human error and variability, providing a standardized approach to identifying pediatric dental conditions across a range of radiographs. Moreover, this automated text generation significantly reduced the time clinicians spent interpreting images, allowing for quicker, more informed decisions regarding diagnosis and treatment. Integrating ChatGPT into the classification workflow helped enhance overall productivity in pediatric dental care, offering support in identifying diseases such as cavities, malocclusions, or developmental abnormalities.

Regarding the impact of resolution reduction on image interpretation, the decreased resolution did not adversely affect ChatGPT’s ability to generate relevant and accurate descriptions for classification. Although high-resolution images offer more detailed visual information, the 25-fold reduction preserved the most important features of pediatric dental diseases, such as signs of tooth decay, alignment issues, and bone abnormalities, which remain visible even at lower resolutions. Since ChatGPT is focused on recognizing and describing these broader patterns, it could still effectively generate descriptions based on the key characteristics of the conditions. However, finer details, such as subtle early signs of disease, may be harder to detect in lower-resolution images. Nonetheless, this trade-off in resolution was justified by the improved processing speed and efficiency, and the reduced-resolution images still supported accurate disease classification for most common pediatric dental conditions.

Table 1 shows the text descriptions obtained from the ChatGPT for eight corresponding dental panoramic radiographs shown in Figure 1.

**Table 1:**
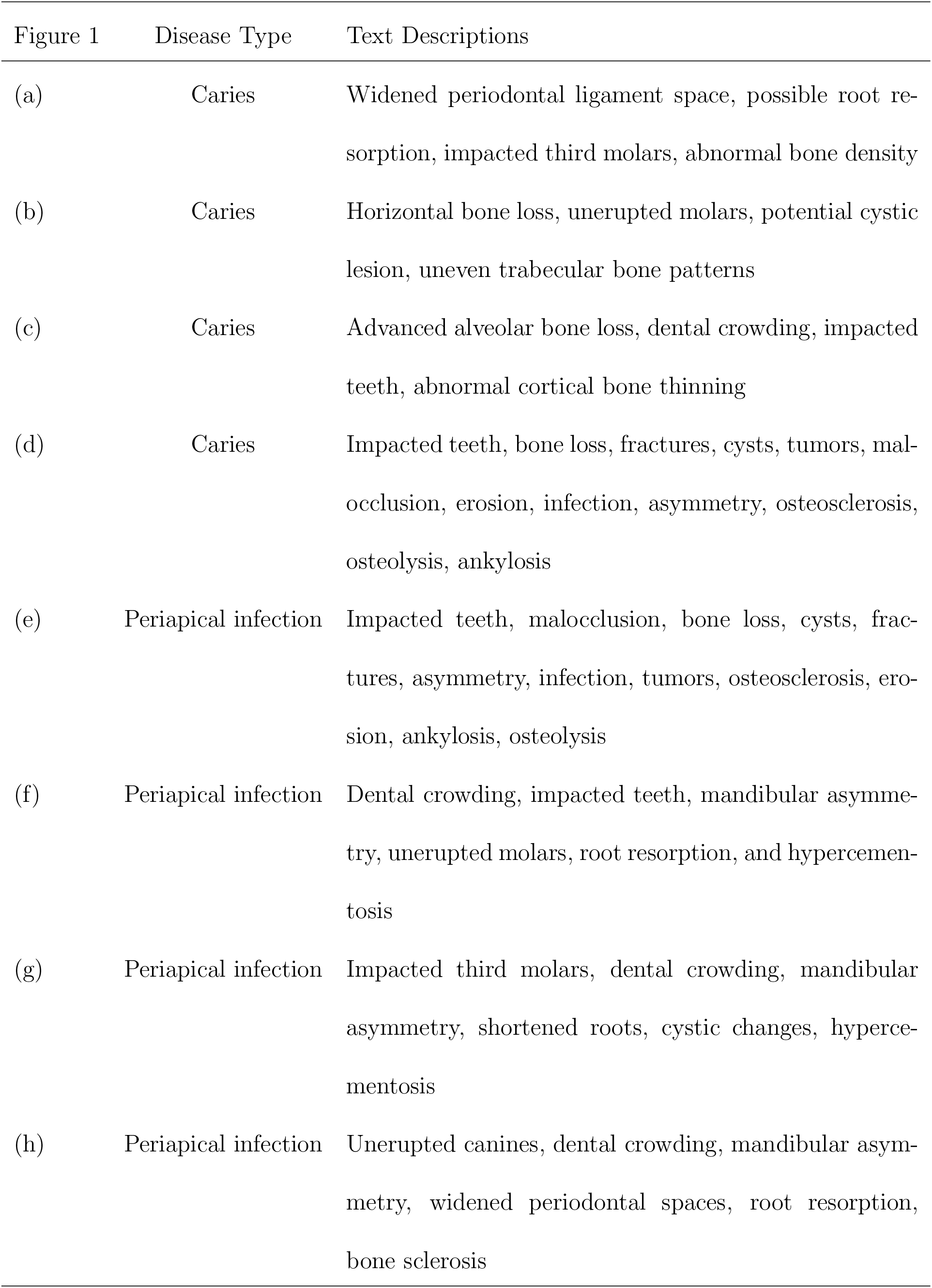
AI-generated descriptions of pediatric dental pathologies on panoramic radiographs using ChatGPT, as shown in Figure 1.

| Figure 1 | Disease Type | Text Descriptions |
| --- | --- | --- |
| (a) | Caries | Widened periodontal ligament space, possible root resorption, impacted third molars, abnormal bone density |
| (b) | Caries | Horizontal bone loss, unerupted molars, potential cystic lesion, uneven trabecular bone patterns |
| (c) | Caries | Advanced alveolar bone loss, dental crowding, impacted teeth, abnormal cortical bone thinning |
| (d) | Caries | Impacted teeth, bone loss, fractures, cysts, tumors, malocclusion, erosion, infection, asymmetry, osteosclerosis, osteolysis, ankylosis |
| (e) | Periapical infection | Impacted teeth, malocclusion, bone loss, cysts, fractures, asymmetry, infection, tumors, osteosclerosis, erosion, ankylosis, osteolysis |
| (f) | Periapical infection | Dental crowding, impacted teeth, mandibular asymmetry, unerupted molars, root resorption, and hypercementosis |
| (g) | Periapical infection | Impacted third molars, dental crowding, mandibular asymmetry, shortened roots, cystic changes, hypercementosis |
| (h) | Periapical infection | Unerupted canines, dental crowding, mandibular asymmetry, widened periodontal spaces, root resorption, bone sclerosis |

## 3 Methods

### 3.1 1D-CNN for Classification

A 1D convolutional neural network (1D-CNN) [52, 53] is designed for processing one-dimensional sequential data, such as time series or signals. The network consists of several key components: convolutional layers, activation functions, pooling layers, fully connected layers, and an output layer with a softmax function for classification.

Let 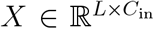 be the input sequence, where *L* is the sequence length, and *C*_in_ is the number of input channels. A 1D convolution operation with kernel size *k* and stride *s* is defined as:

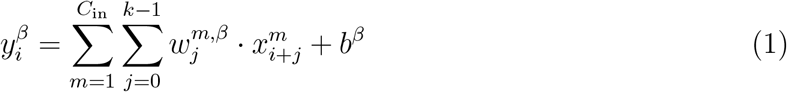

where 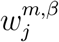 is the convolution kernel for input channel *m* and output channel *β, b*^*β*^ is the bias term, 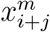 is the input at position *i* + *j* for channel *m*, and 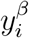 is the feature map output at position *i* for channel *β*.

The output feature map has dimensions (*L*^′^, *C*_out_), defined as

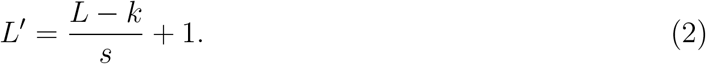

A non-linear activation function *σ*(·) known as the ReLU activation is applied element-wise:

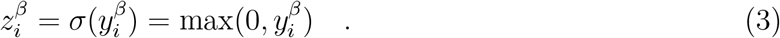

To reduce dimensionality, max pooling with pooling size *p* is applied as follows.

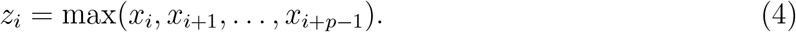

The output length after pooling is

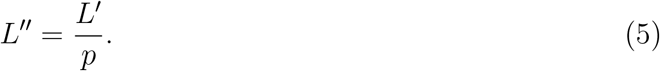

The extracted features are flattened and passed through a fully connected layer:

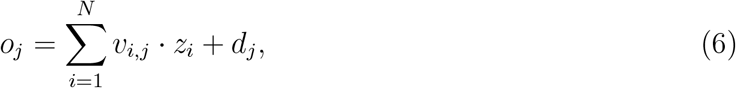

where *v*_*i,j*_ are weights, *d*_*j*_ are biases, and *N* is the number of flattened features.

For classification with *C* classes, the final layer applies the softmax function:

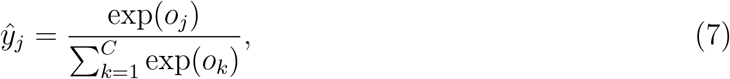

where *ŷ*_*j*_ is the predicted probability for class *j*.

Given the true class label *y*_*j*_ (one-hot encoded), the loss function is defined in terms of the Shannon entropy as

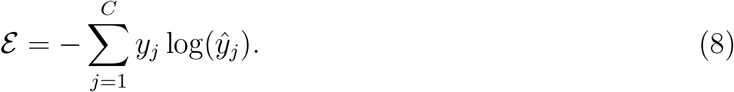

A 1D-CNN assigns a feature to a class through the learned feature representations and the classification layer. The decision-making process follows these steps:

1. Extracted features from convolutional and pooling layers form a feature vector **z** ∈ ℝ^*N*^.
2. The fully connected layer computes a set of logits **o** ∈ ℝ^*C*^, where *o*_*j*_ represents the unnormalized score for class *j*.
3. The softmax function converts logits into probabilities *ŷ*_*j*_ described in Eq. (7).
4. The predicted class *c*^*^ is assigned as:

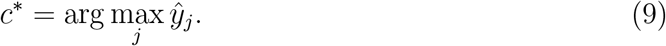
5. The final classification decision is made based on *c*^*^, which corresponds to the class with the highest probability.

### 3.2 Word Encoding

Let *𝒟* = {*D*_1_, *D*_2_, …, *D*_*Q*_} be a collection of *Q* documents, where each document *D*_*i*_ is a sequence of tokens (words). Each document *D*_*i*_ consists of *T*_*i*_ tokens as

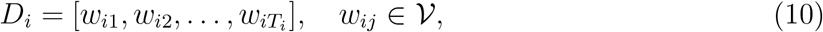

where *𝒱* is the vocabulary (the set of all unique tokens across all documents).

The vocabulary is defined as the union of all tokens in all documents:

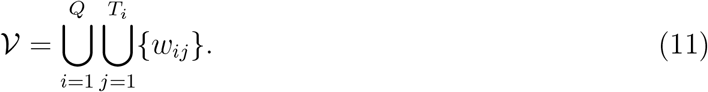

Let | *𝒱*| = *V* be the vocabulary size. A bijective function is defined as

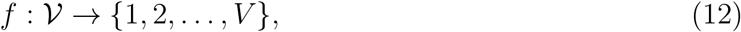

such that each word *w* ∈ *𝒱* is mapped to a unique index *i* ∈ ℤ_≥1_:

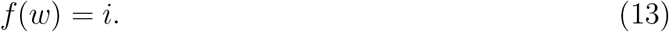

The inverse mapping is

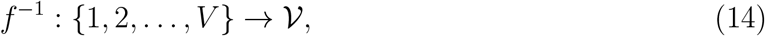

which is used to retrieve the word corresponding to an index.

Each document *D*_*i*_ is encoded as a sequence of integer indices as

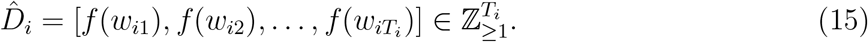

For any token *w* ∉*𝒱*, a special unknown token is assigned by

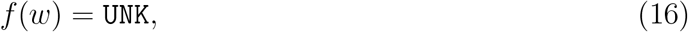

where `UNK` is a reserved index (e.g., 0) for unseen words.

### 3.3 Gray-Level Co-Occurrence Matrix of a Fuzzy Recurrence Plot

Let *I*(*x, y*) denote the grayscale intensity at pixel (*x, y*) in a 2D image of size *ℳ* × *𝒩*, with *ℒ* possible gray levels:

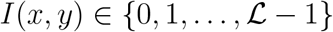

The gray-level co-occurrence matrix (GLCM), denoted as *G*_*d,θ*_(*i, j*), is defined as the number of times two pixels with intensity values *i* and *j* occur in the image, separated by a spatial offset vector (*dx, dy*), corresponding to a distance *d* and direction *θ*.

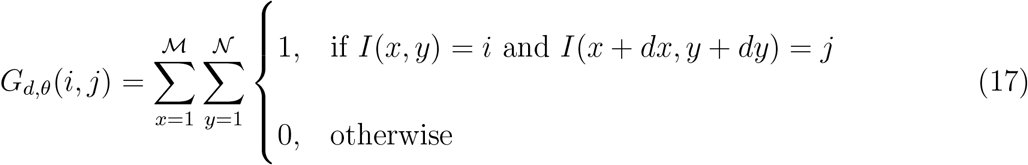

The offset (*dx, dy*) is determined by the direction *θ* and distance *h*. Common directions include: *θ* = 0^°^ → (*dx, dy*) = (*h*, 0), *θ* = 45^°^ → (*dx, dy*) = (*h*, −*h*), *θ* = 90^°^ → (*dx, dy*) = (0, −*h*), and *θ* = 135^°^ → (*dx, dy*) = (−*h*, −*h*).

The resulting GLCM is a matrix of size *ℒ* × *ℒ*, where each element (*i, j*) counts how often a pixel with intensity *i* is found adjacent to a pixel with intensity *j* at the specified offset.

In this study, the image *I* is represented with a fuzzy recurrence plot (FRP) [55, 56], which models the nonlinear dynamics of the vectorized panoramic radiograph.

Let **r** = (*r*_1_, *r*_2_, …, *r*_*T*_) denote the vectorized pixel intensities of a panoramic radiograph. The phase space representation of **r**, denoted as Ω, is constructed using time-delay embedding:

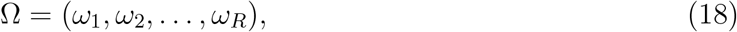

in which the number of state vectors *R* is given by:

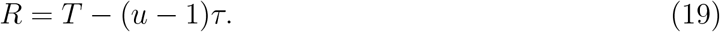

where *u* is the embedding dimension, *τ* is the time delay, and each state vector *ω*_*i*_ is defined as:

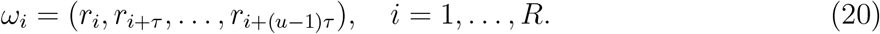

The FRP matrix **F** quantifies the degree of recurrence among state vectors in the phase space and is constructed by applying the fuzzy *c*-means (FCM) clustering algorithm [57] and fuzzy relations [58] to Ω. The recurrence matrix is defined as:

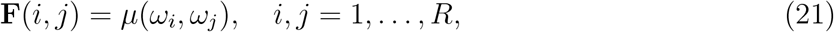

where *µ*(*ω*_*i*_, *ω*_*j*_) denotes the degree of similarity between state vectors *ω*_*i*_ and *ω*_*j*_, determined by the following properties:

1. *Self-similarity:*

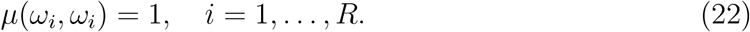
2. *Symmetry:*

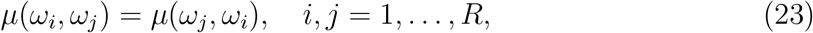

where 0 *< µ*(*ω*_*i*_, *ω*_*j*_) *<* 1 represents the degree of similarity between *ω*_*i*_ and *ω*_*j*_.
3. *Transitivity:*

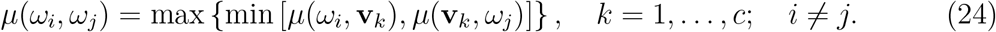

The similarity values *µ*(*ω*_*i*_, **v**_*k*_) are computed using the fuzzy *c*-means (FCM) algorithm [57], which is a clustering method that partitions a dataset into *c* fuzzy clusters by minimizing an objective function based on membership degrees.

Given a dataset Ω = (*ω*_1_, *ω*_2_, …, *ω*_*R*_}, the FCM algorithm seeks to find cluster centers **V** = {**v**_1_, **v**_2_, …, **v**_*c*_} and a fuzzy partition matrix **U** = [*µ*_*ik*_] ∈ ℝ^*c*×*R*^, where *µ*_*ik*_ represents the degree of membership of *ω*_*k*_ in cluster **v**_*i*_.

The objective function to be minimized is:

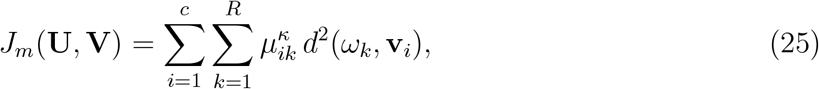

where *κ >* 1, which is commonly taken as 2 and used in this study, is the fuzziness parameter that controls the degree of fuzziness in the clustering, *d*^2^(**x**_*k*_, **v**_*i*_) = ∥*ω*_*k*_ − **v**_*i*_∥^2^ is the squared Euclidean distance between the data point *ω*_*k*_ and the cluster center **v**_*i*_.

The membership values *µ*_*ik*_ and cluster centers **v**_*i*_ are updated iteratively using the following update equations:

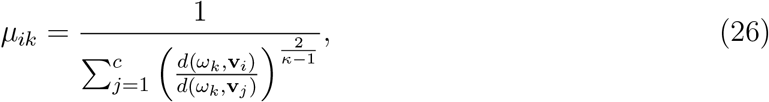

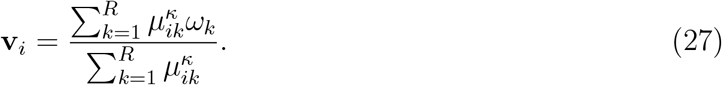

The algorithm iterates until the partition matrix **U** converges, typically when the maximum change in membership values between consecutive iterations falls below a predefined threshold *ϵ*:

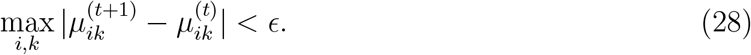

The FCM algorithm provides soft clustering, where each data point can belong to multiple clusters with different degrees of membership rather than being assigned to a single cluster.

### 3.4 Integration of text and image features

The text descriptions of panoramic radiographs are transformed into sequences of numeric indices or word vectors, making them suitable for input into deep learning models. Each narrative is converted into a sequence of numeric indices corresponding to words in the encoding’s vocabulary, which is built using MATLAB’s Text Analytics Toolbox (https://mathworks.com/products/text-analytics.html). This approach is particularly useful for models that process word sequences as categorical inputs.

To ensure uniform input lengths for batch processing, shorter sequences are left-padded, which is essential for models requiring fixed-length inputs. Additionally, a key feature of text encoding is its handling of out-of-vocabulary words. By default, tokens not present in the embedding’s vocabulary are discarded; however, an alternative handling method can be specified using the “UnknownWord” option, allowing these words to be mapped to a predefined vector representation.

The word vectors extracted from the text descriptions of the panoramic radiographs are concatenated with the vectorized GLCMs, forming a unified feature representation. This integration of linguistic and structural information enables a multimodal approach, where both textual and visual features contribute to learning meaningful patterns. These combined features serve as input to the 1D-CNN, which processes the sequential data to extract discriminative representations. Together, these two types of features constitute a vision-language model for deep learning, allowing the network to leverage complementary information from both modalities for improved classification performance.

### 3.5 Implementation

The 1D-CNN architecture was specifically designed for classification tasks using concatenated features derived from the vectorized GLCMs of the FRPs and their corresponding text encodings. For FRP construction, the embedding dimension was set to 3, the time delay to 1, and the number of clusters to 3. In generating the GLCMs from the FRPs, the number of gray levels was fixed at 4, and the spatial offset between the pixel of interest and its neighbor was set at 0^°^.

The deep network has the input size as 1, which corresponds to the channel dimension of the input integer sequence. To process the input data, a word embedding layer was applied, where the dimension of the embedding was set to 50, transforming each word into a fixed-length vector representation.

Following the embedding layer, the architecture incorporates multiple blocks designed to capture features at varying *n*-gram lengths, specifically for *n* = 2, 3, 4, and 5. Each block is composed of a series of layers: 1) a 1D convolutional layer, which applies convolutions over the input sequence to capture local patterns corresponding to the specified *n*-gram length; 2) a batch normalization layer, which normalizes the activations of the convolutional layer to improve training stability; 3) a ReLU activation layer, which introduces non-linearity to the model, enabling it to learn more complex patterns. 4) a dropout layer, with a rate of 0.2, to reduce overfitting by randomly setting a fraction of input units to zero during training; and 5) a global max pooling layer, which reduces the dimensionality of the output by selecting the maximum value from each feature map.

Each block uses 100 convolutional filters, with the filter size matching the *n*-gram length being modeled. The outputs from each of these blocks are concatenated together using a concatenation layer, which allows the model to simultaneously learn features from different *n*-gram lengths and capture a broader range of patterns in the text.

The final part of the architecture includes a fully connected (dense) layer, which processes the concatenated feature maps from the previous layers, followed by a softmax layer that outputs the class predictions. This softmax layer ensures that the network performs multi-class classification by assigning probabilities to each class. The overall structure of the network allows it to handle multiple *n*-gram lengths efficiently by connecting each convolutional block to the word embedding layer and merging their outputs at the concatenation stage.

The network was trained using the Adam optimizer, with a mini-batch size of 128, to ensure effective and efficient training. Cross-entropy loss was employed as the optimization objective, which is well-suited for classification tasks. During training, a separate validation dataset was used to evaluate the model’s performance at each epoch. The model that exhibited the lowest validation loss was saved for future use, ensuring that it generalizes well to unseen data.

Figure 2 presents a graphical procedure for constructing the proposed deep learning-based VLM, illustrating the key components, data flow, and integration strategy between visual and textual modalities.

**Figure 2:**
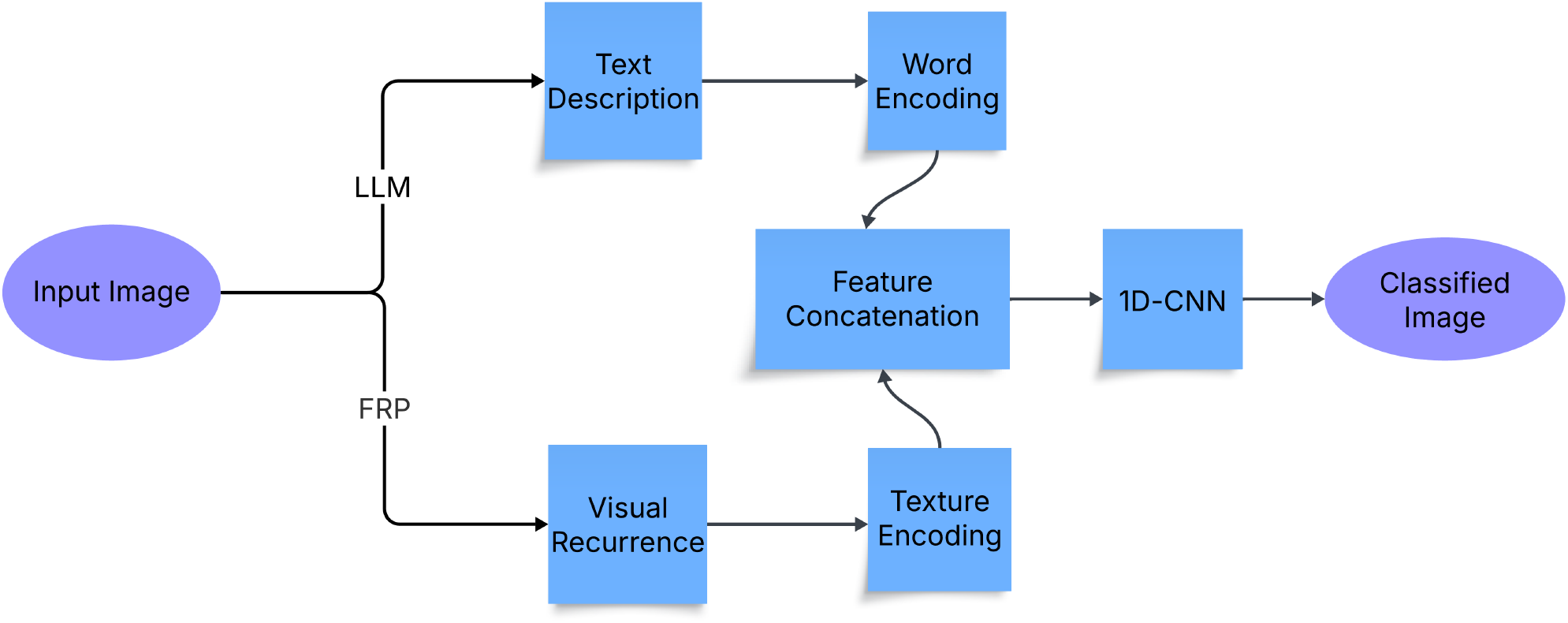
Graphical procedure for diagnosing pediatric dental diseases on dental panoramic radiographs using the proposed deep learning VL model.

## 4 Results

Following the previous study [47], this study addresses a binary classification problem aimed at distinguishing between caries and periapical infections. Classification outcomes are defined using standard terminology: a true positive (TP) occurs when a periapical infection is correctly identified as such, while a false positive (FP) arises when a case of caries is incorrectly labeled as a periapical infection. Conversely, a true negative (TN) denotes a caries case correctly classified, and a false negative (FN) indicates a periapical infection misclassified as caries.

To evaluate model performance, several key metrics are employed. Accuracy (ACC) quantifies the overall proportion of correct predictions, encompassing both TP and TN, relative to the total number of cases. Sensitivity (SEN), also known as recall, measures the model’s ability to correctly detect periapical infections, whereas specificity (SPE) assesses its capacity to correctly identify caries. Precision (PRE) indicates the proportion of instances predicted as periapical infections that are actually correct, offering insight into the reliability of positive predictions. The F1 score combines precision and sensitivity into a single metric by computing their harmonic mean. This score is particularly valuable in assessing the balance between false positives and false negatives, with a focus on accurately identifying periapical infections. Finally, the area under the receiver operating characteristic curve (AUC) provides a comprehensive measure of the classifier’s discriminative ability. It captures the trade-off between sensitivity and specificity across varying decision thresholds, with higher AUC values indicating stronger overall performance and robustness in distinguishing between the two dental conditions.

The dataset was partitioned using a holdout strategy, with 90% allocated for training and the remaining 10% reserved for validation. Table 2 presents the comparative performance of previously reported and proposed AI models, while Table 3 summarizes their performance across key evaluation metrics.

**Table 2:** Comparison of performance metrics across different AI models.

| Model | ACC (%) | SEN (%) | SPE (%) | PRE (%) | F1 | AUC |
| --- | --- | --- | --- | --- | --- | --- |
| Pretrained CNN |  |  |  |  |  |  |
| SqueezeNet [47] | 50.00 | 46.67 | 53.33 | 54.17 | 0.49 | 0.62 |
| GoogLeNet [47] | 56.67 | 66.67 | 46.67 | 58.67 | 0.61 | 0.62 |
| AlexNet [47] | 66.67 | 73.33 | 60.00 | 63.33 | 0.67 | 0.73 |
| Deep learning language models |  |  |  |  |  |  |
| LSTM [47] | 56.67 | 75.00 | 41.67 | 55.56 | 0.62 | 0.72 |
| BERT [47] | 76.67 | 83.33 | 66.67 | 83.33 | 0.82 | 0.73 |
| 1D-CNN [47] | 84.00 | 86.67 | 86.67 | 86.67 | 0.84 | 0.93 |
| Proposed deep learning vision-language model |  |  |  |  |  |  |
| VLM | 90.00 | 91.67 | 83.33 | 91.67 | 0.90 | 0.96 |

**Table 3:** Comparative model performance across key metrics.

| Metric | Best Model | Key Comparison |
| --- | --- | --- |
| Accuracy | VLM | Substantially better than 1D-CNN and BERT |
| Sensitivity | VLM | Slightly higher than 1D-CNN and BERT |
| Specificity | 1D-CNN | VLM slightly lower , but with better overall balance |
| Precision | VLM | Tied with its own sensitivity; highest among all models |
| F1 Score | VLM | Outperforms 1D-CNN and BERT |
| AUC | VLM | Indicates superior discriminative ability over 1D-CNN |

Figure 3 illustrates the transformation of a panoramic radiograph into structural features for integration with corresponding text-encoded information. The original image is simultaneously processed in two parallel streams: one generates a textual description using a large language model (ChatGPT), while the other extracts visual recurrence patterns using the FRP method. The textual and visual data are then encoded using the word embedding and GLCM techniques, respectively, resulting in two distinct feature sets. These features are subsequently concatenated and used to train and validate the 1D-CNN classification model.

**Figure 3:**
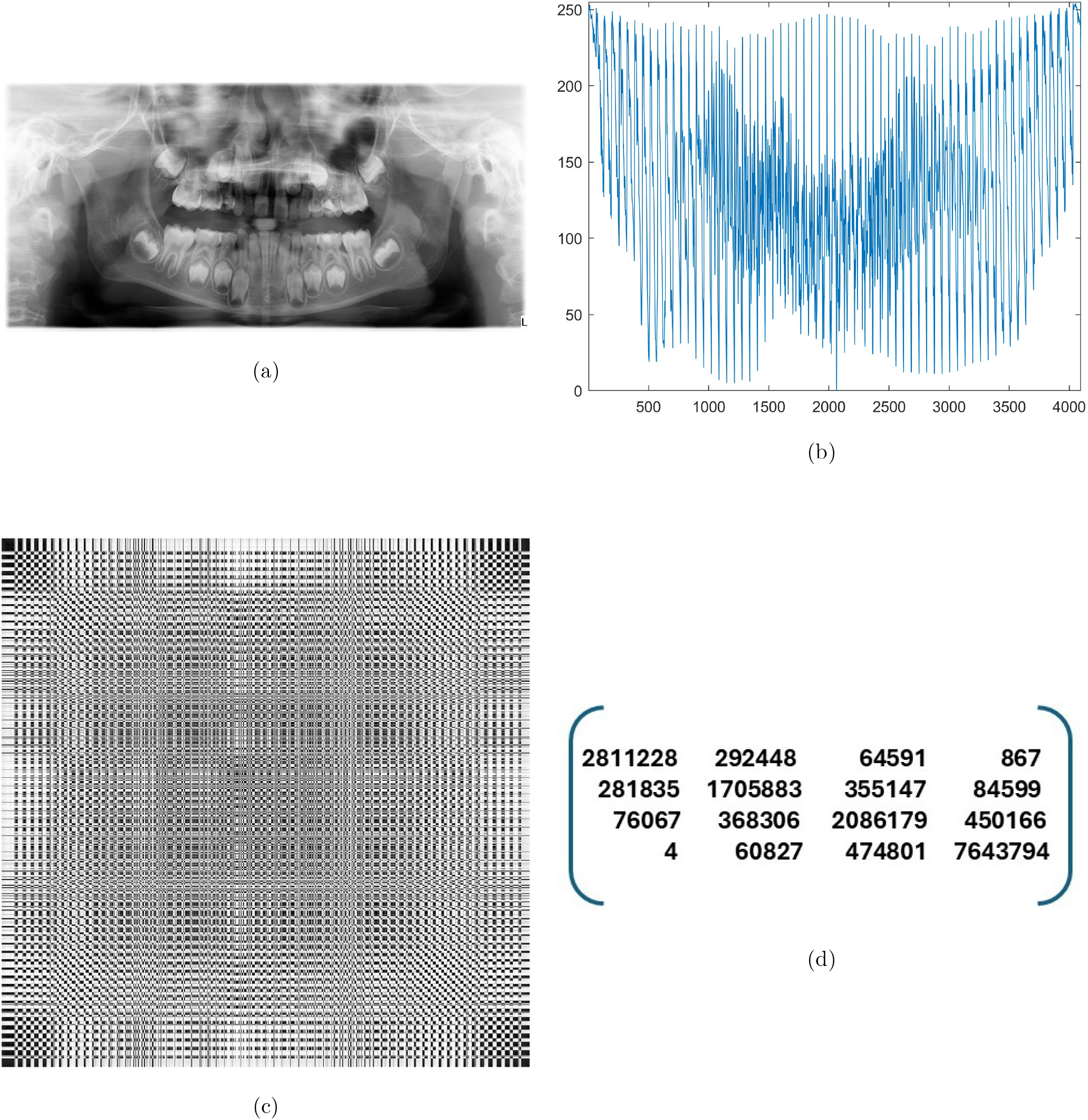
Example of the sequential transformation of an image to texture encoding: 1000×471 dental panoramic radiograph (a), vectorization of the panoramic radiograph resized to 64 × 64 pixels (b), FRP of resized OPG (c), and GLCM with 4 gray levels of the FRP (d).

As another example of getting insight into the proposed model’s learning behavior and stability under different initialization conditions., Figure 4 presents the training and validation progress of the deep learning vision-language model using two different random seeds. In Figure 4(a) and (b), the model demonstrates a consistent learning trajectory. The training loss decreases with small fluctuations over epochs, accompanied by a parallel decline in validation loss. Similarly, both training and validation accuracy improve in a coordinated manner, indicating that the model is not only learning effectively but also generalizing well to unseen data. The close alignment between training and validation curves suggests that the model trained with this particular random seed maintains a strong generalization capacity and is relatively robust against overfitting. This pattern reflects a stable optimization process, where the model consistently learns meaningful features from the input data across epochs.

**Figure 4:**
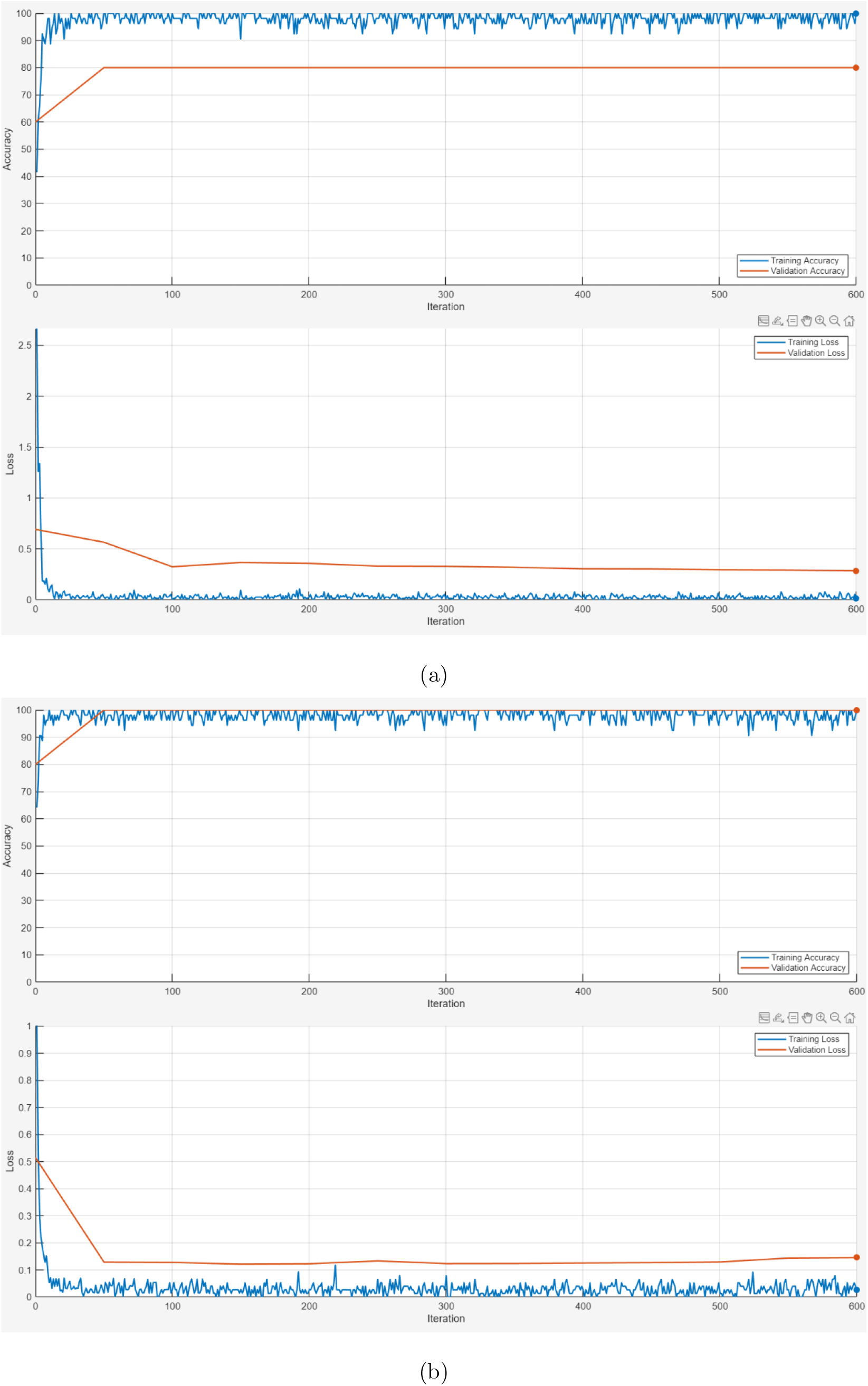
Training and validation progresses of the proposed deep learning VL model using two different random generation seeds, as shown in (a) and (b).

## 5 Discussion

The models are categorized into three groups: pretrained CNNs, deep learning language models, and the proposed VLM. Among the pretrained CNNs, AlexNet outperforms SqueezeNet and GoogLeNet across all metrics, achieving an accuracy of 66.67%, sensitivity of 73.33%, and specificity of 60.00%. However, even AlexNet shows limited precision (63.33%) and a modest F1 score (0.67), indicating a suboptimal balance between the detection of periapical infections and avoidance of false positives. SqueezeNet, with an accuracy of 50% and a nearly random classification profile, performs the worst, while GoogLeNet slightly improves upon this but remains imbalanced, with higher sensitivity (66.67%) at the cost of specificity (46.67%). Overall, the pretrained CNNs demonstrate limitations in generalizing effectively to this diagnostic task, likely due to their reliance on visual features without contextual linguistic support.

In contrast, the deep learning language models present a noticeable performance improvement. LSTM achieves a high sensitivity (75.00%) but suffers from low specificity (41.67%), leading to poor discrimination of caries cases. Its F1 score of 0.62 and moderate AUC (0.72) reflect this imbalance. BERT, leveraging transformer-based language understanding, demonstrates a stronger balance, with 83.33% sensitivity, 66.67% specificity, and a high F1 score of 0.82. It also achieves a significantly improved precision of 83.33%, highlighting its capability in accurately identifying periapical infections. 1D-CNN, though structurally simpler, outperforms both LSTM and BERT with an accuracy of 84.00% and equal sensitivity and specificity of 86.67%. The F1 score (0.84) and AUC (0.93) suggest that th7is model maintains a well-calibrated performance across both diagnostic classes.

The proposed vision-language model (VLM), which integrates both visual and linguistic features, delivers the highest performance across all metrics. It achieves an accuracy of 90.00%, sensitivity of 91.67%, and precision also at 91.67%, indicating not only its robustness in correctly identifying periapical infections but also in minimizing false positives. Specificity, at 83.33%, is slightly lower than that of the 1D-CNN but still demonstrates excellent ability in correctly identifying caries. The F1 score of 0.90 underscores the model’s balanced performance, and the AUC of 0.96 confirms its superior discriminative capacity. This high level of performance is likely attributed to the model’s multimodal architecture, which enables a more comprehensive understanding of the diagnostic task by contextualizing visual cues with language-based information.

While pretrained CNNs and standalone language models offer varying levels of success, the proposed VLM stands out for its consistent and superior performance. Its ability to combine modalities appears to significantly enhance its diagnostic accuracy and reliability, making it a promising candidate for clinical deployment in dental radiographic analysis focused on distinguishing between periapical infections and caries.

While the proposed VLM demonstrates promising performance in diagnosing pediatric dental diseases, this study is limited by the relatively small dataset focused only on two disease classes: caries and periapical infections. The generalizability of the model to broader clinical settings and more diverse diagnostic categories remains to be validated. Additionally, the reliance on automatically generated text descriptions may introduce variability in semantic accuracy. Future research should explore the integration of larger, more heterogeneous datasets, include additional diagnostic categories, and evaluate model performance in real-world clinical workflows. Further enhancement of the language component through domain-specific fine-tuning of large language models may also improve interpretability and diagnostic precision.

## 6 Conclusion

This study presents a novel deep learning vision-language model for the classification of pediatric dental diseases, specifically focusing on distinguishing between caries and periapical infections. By integrating visual texture features derived from FRPs and structural representations using GLCMs with text-based features generated by a large language model, the proposed approach demonstrates superior performance compared to traditional CNNs and standalone language models. The model achieved high accuracy, sensitivity, and precision, highlighting the effectiveness of multimodal feature fusion in enhancing diagnostic capability. These results underscore the potential of vision-language models in advancing AI-assisted diagnosis in dental radiology, paving the way for more interpretable, efficient, and clinically relevant decision support systems.

Beyond its immediate diagnostic performance, the proposed framework exemplifies the potential of multimodal AI systems to bridge the gap between visual data and clinical narratives, enhancing the interpretability and reliability of automated decision-making in healthcare. As dental diagnostics increasingly incorporate AI tools, models that effectively combine image and text information can offer more context-aware and explainable outputs. Future expansions of this work may include multi-class classification across a wider range of dental pathologies, real-time integration into clinical software, and further validation through prospective studies in diverse clinical settings. Such advancements will be critical for translating AI-driven models from proof-of-concept research into practical tools that support clinicians and improve patient outcomes.

## Author contribution

TDP contributed to conception, technical design, computer coding and implementation, data interpretation, and writing the manuscript.

## Data availability

The children’s dental panoramic radiographs dataset is publicly available at https://doi.org/10.6084/m9.figshare.c.6317013.v1. AI-generated text descriptions corresponding to these radiographs are available on the author’s personal website (https://sites.google.com/view/tuan-d-pham/codes), under the title “Image-to-text classification of dental diseases”.

## Software availability

MATLAB codes implemented in this study are available at the author’s personal website (https://sites.google.com/view/tuan-d-pham/codes), under the title “VLM for diagnosing dental diseases”.

